# Comparative transcriptomic analyses of peripheral blood mononuclear cells of patients with non-pneumonia and severe pneumonia at 1 year-Long-COVID-19

**DOI:** 10.1101/2023.12.12.23299822

**Authors:** Ozgecan Kayalar, Pelin Duru Cetinkaya, Vahap Eldem, Serap Argun Baris, Nurdan Koktürk, Selim Can Kuralay, Hadi Rajabi, Nur Konyalilar, Deniz Mortazavi, Seval Kubra Korkunc, Sinem Erkan, Gizem Tuşe Aksoy, Gul Eyikudamaci, Pelin Pinar Deniz, Oya Baydar Toprak, Pinar Yildiz Gulhan, Gulseren Sagcan, Neslihan Kose, Aysegul Tomruk Erdem, Fusun Fakili, Onder Ozturk, Ilknur Basyigit, Hasim Boyaci, Emel Azak, Tansu Ulukavak Ciftci, Ipek Kivilcim Oguzulgen, Hasan Selcuk Ozger, Pinar Aysert Yildiz, Ismail Hanta, Ozlem Ataoglu, Merve Ercelik, Caglar Cuhadaroglu, Hacer Kuzu Okur, Muge Meltem Tor, Esra Nurlu Temel, Seval Kul, Yıldız Tutuncu, Oya Itil, Hasan Bayram

## Abstract

Long-COVID-19 manifests as a multisystemic condition with varied symptoms lingering beyond three weeks of acute SARS-CoV-2 infection, though its underlying mechanisms remain elusive. Aiming to decipher the long-term molecular impacts of COVID-19, we conducted a transcriptomic analysis on PBMCs from 1-year post-covid patients, including individuals without pneumonia (NP, n=10), those with severe pneumonia (SP, n=11), and healthy controls (C, n=13). Our extensive RNA sequencing revealed 4843 differentially expressed genes (DEGs) and 1056 differentially expressed long non-coding RNAs (DElncRNAs) in “C vs NP,” 1651 DEGs and 577 DElncRNAs in “C vs SP,” 954 DEGs and 148 DElncRNAs in “NP vs SP,” with 291 DEGs and 70 DElncRNAs shared across all groups. We identified 14 hub genes from 291 DEGs, with functional enrichment analysis showing upregulated DEGs mainly linked to inflammation and osteoclast differentiation, and downregulated DEGs to viral infections and immune responses. These hub genes play central roles in inflammatory and immune processes and are significantly associated with pneumonitis and diverse lung diseases. Investigations revealed unique immune cell signatures across DEG categories, associating upregulated DEGs with neutrophils and monocytes, and downregulated DEGs with CD4 memory effector T cells. Analysis of 14 hub genes showed notable upregulation in the no pneumonia group versus healthy controls, displaying complex patterns in the severe pneumonia group. Our study uncovered potential idiopathic pulmonary fibrosis signals in Long-COVID-19 patients’ PBMC transcriptome, highlighting the urgency for thorough monitoring and extended research to understand COVID-19’s lasting effects. This study sheds light on COVID-19’s transcriptomic changes and potential lasting effects, guiding future research and therapeutic approaches for Long-COVID-19.

## 1. Introduction

SARS-CoV-2, a highly contagious respiratory virus that is the source of the continuing worldwide pandemic, is the cause of the coronavirus disease 2019 (COVID-19). Common symptoms of COVID-19 include fever, cough, headache, lethargy, myalgia, diarrhoea, and anosmia in previously healthy people. COVID-19 often manifests as an asymptomatic or mild to moderate respiratory infection [1, 2]. Additionally, COVID-19 in patients with pre-existing co-morbidities such as obesity, respiratory, cardiovascular, and renal illnesses can swiftly progress into a serious, life-threatening condition needing immediate critical care support [3, 4]. According to the World Health Organization (WHO), there were 6,897,025 fatalities and 762,791,152 confirmed cases of COVID-19 worldwide as of 12 April 2023 (WHO, 2023).

The early acute stages of COVID-19 have been the subject of numerous studies, but studies of the long-term consequences of the disease have received much less attention, and still little is known. In the past two years, studies have reported that 30-70% of recovered individuals who had mild to severe COVID-19 struggling with prolonged symptoms for the time more than a 1 year after infection[5–10], have been commonly referred to as Long-COVID (CDC, 2021) or post-acute sequelae of COVID-19 (PASC)[11]. Long-COVID-19 is defined as a multisystemic condition with severe symptoms for at least 3 weeks or longer after the first negative PCR test following the acute course of SARS-CoV-2 infection [12]. Most cases are resolved within two to four weeks of the initial symptoms’ appearance; however, after the initial infection, early studies have showed that these persistent symptoms in some cases may appear four weeks to less than a year later [13, 14]. The most common symptoms observed in the Long-COVID-19 studies include fatigue, headache, attention disorder, memory loss, hair loss, gastrointestinal (GI) distress, dyspnoea, anosmia, shortness of breath, pneumonia, and other symptoms[9, 15, 16].

Recent studies have revealed that Long-COVID-19 is associated with the features of the severity of the disease [7, 17] including the presence of chronic pulmonary fibrosis, pneumonia at initial diagnosis, dyspnoea, and emergency service admission in the Long-COVID-19 period [8]. Long-COVID-19 has been thought to be related to autoimmune mechanisms, unresolved viral fragments, and the presence of high-titre SARS-CoV-2 IgG response induced by the natural infection [7, 18, 19]. Additionally, although some infected people experience long-term radiological abnormalities linked to pulmonary function impairment [20–23], the underlying pathogenic pathways are poorly understood. Besides, it is unclear why some patients experience complete clinical, physiological, and radiological recovery while others have a more dangerous outcome that includes persisting interstitial lung alterations and related pulmonary function impairment. It is still unknown whether the COVID-19 sequelae that are still present in the lungs are a result of the lingering effects of the initial increase in the inflammatory response or the activation of alternate pathways after the acute illness has subsided. To address these knowledge gaps, we carried out unbiased next-generation RNA sequencing from peripheral blood mononuclear cells (PBMCs) of the patients with the absence of pneumonia and presence of severe pneumonia in 1-year recovery after COVID-19 and the healthy controls without SARS-CoV-2 infection.

## 2. Material and methods

### Ethical Statement

This study followed the principles of the Declaration of Helsinki and was approved by the institutional ethics committee of Çukurova University School of Medicine (Approval number: 356/22.05.2021). Informed consent was obtained from the patients and the healthy volunteers for the collection of blood samples.

### Data Availability

FASTQ sequences of the PBMC samples have been deposited in NCBI Short Read Archive (SRA) under BioProject PRJNA895325.

### Study population

This study was conducted on patients who participated in the previously published TURCOVID study [8, 24]. During the first wave of the COVID-19 pandemic (between 11 March and 18 July 2020), 1 500 patients over the age of 18, who were monitored and treated because of COVID-19 were included in the targeted trial population in the multi-centre TTS-TURCOVID-19 registry cohort. A total of 831 patients were enrolled in the trial at 13 of the 26 locations (11 university hospitals, 2 sizable tertiary institutions, and 1 private hospital). A standard questionnaire was applied to current patients in the cohort over the phone after receiving written informed consent. Of the cohort of 831 patients, 272 (32.7%) could not be reached, 48 (5.8%) refused to participate in the study, 69 (8.3%) were excluded due to death, and the remaining 442 patients were included (Form-1). Retrospective data entry was performed over the recorded files, and an analysis of the medical records of 442 patients, who could be reached by phone, was applied (Form-2). One year later, 138 patients from 11 centres, who agreed to participate in the study and filled out a signed informed consent form, were called for a follow up check. A routine evaluation (clinical, laboratory and radiological) of the patients was performed (Form-3). From these cases, 27 were randomized into two groups using the computer program at https://www.medcalc.org/. Two groups were formed: one consisting of 13 cases (male: 8; female: 5) without pneumonia in their radiology findings after experiencing COVID-19, and the other consisting of 14 cases (male: 10; female: 4) with severe pneumonia. The control group was composed of 13 cases (male: 8; female: 5), who remained uninfected. Age, gender, and smoking status were taken into consideration to eliminate confounding factors in the randomization process. Thus, the cases were divided into three groups: (i) those with no pneumonia following COVID-19 infection, (ii) those with severe pneumonia during COVID-19 infection, and (iii) the healthy control group with no disease (Table 1). Peripheral blood mononuclear cells (PBMC) were isolated from the blood obtained from the patients and stored at −80LC until used.

**Table 1.**
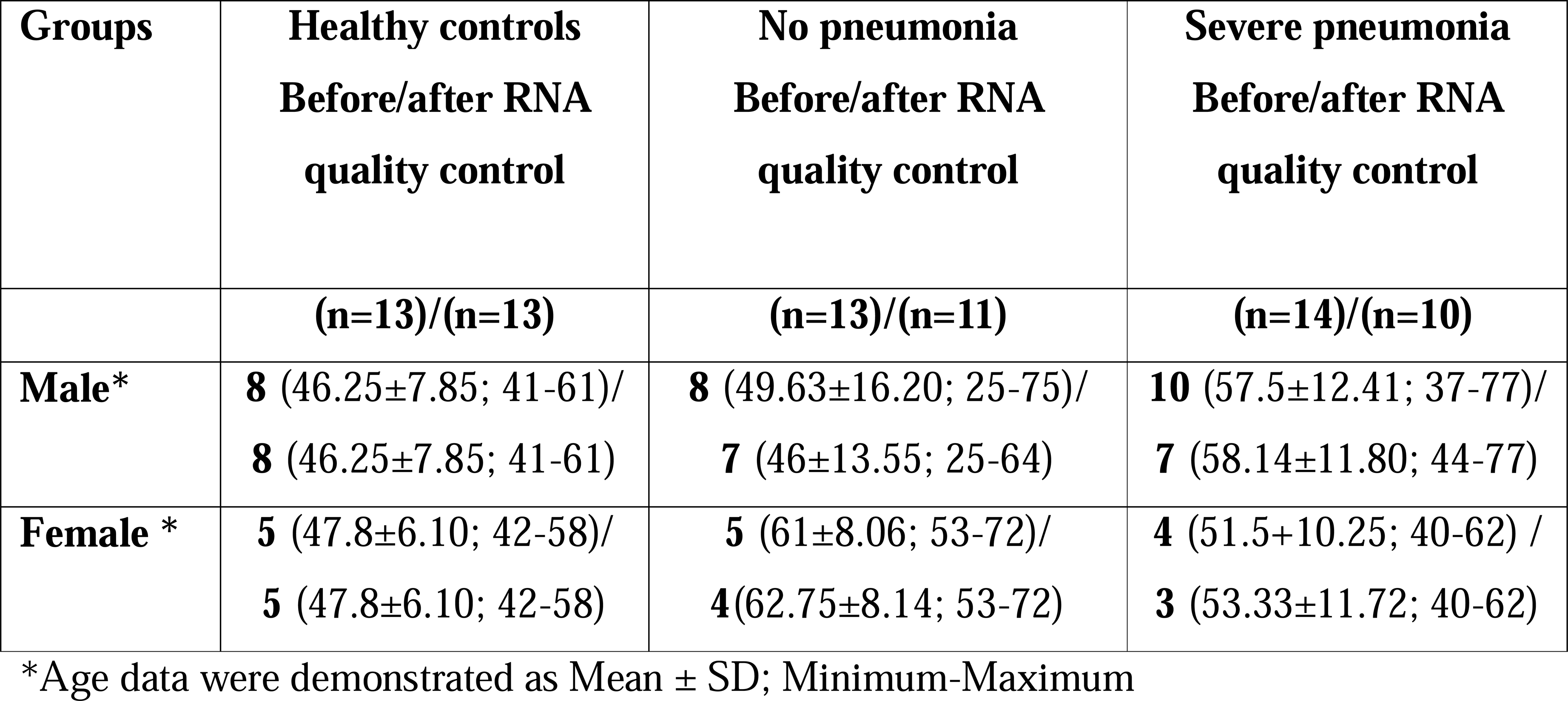
Demographics of study subjects.

| **Groups** | **Healthy controls**<br>**Before/after RNA**<br>**quality control** | **No pneumonia**<br>**Before/after RNA**<br>**quality control** | **Severe pneumonia**<br>**Before/after RNA**<br>**quality control** |
| --- | --- | --- | --- |
|  | **(n=13)/(n=13)** | **(n=13)/(n=11)** | **(n=14)/(n=10)** |
| **Male\*** | **8** (46.25±7.85; 41-61)/<br>**8** (46.25±7.85; 41-61) | **8** (49.63±16.20; 25-75)/<br>**7** (46±13.55; 25-64) | **10** (57.5±12.41; 37-77)/<br>**7** (58.14±11.80; 44-77) |
| **Female \*** | **5** (47.8±6.10; 42-58)/<br>**5** (47.8±6.10; 42-58) | **5** (61±8.06; 53-72)/<br>**4**(62.75±8.14; 53-72) | **4** (51.5+10.25; 40-62) /<br>**3** (53.33±11.72; 40-62) |

\*Age data were demonstrated as Mean ± SD; Minimum-Maximum

### Blood Sample Collection and Isolation of Peripheral Blood Mononuclear Cells (PBMCs)

Approximately 20 ml of venous blood was collected from each participant, and PBMCs were isolated using a solution of Lymphoprep™ (Alere Technologies, Norway) by performing density gradient sedimentation at 2000 rpm for 20 minutes.

### Transcriptome Library Construction and Next-generation RNA Sequencing

Following isolation, the purity and integrity of RNA was assessed using a NanoDrop^TM^ spectrophotometer (Thermo Scientific, Nanodrop 2000c) and RNA Nano 6000 Assay Kit of the Agilent Bioanalyzer 2100 system (Agilent Technologies, CA, USA), respectively. Samples with RNA integrity number values >7.5 were retained for further processing. Before library preparation, mRNA and long non-coding RNAs (lncRNAs) were enriched by MGIEasy rRNA Depletion Kit (MGI Tech, China), and DNase I treatment (NEB) was carried out for complete removal of the DNA according to the manufacturer’s instruction. RNA libraries were constructed from 500 ng RNA using the MGIEasy RNA Library Prep Kit V3.0 protocol (MGI, Shenzen, China) according to the manufacturer’s instruction. Firstly, enriched RNA samples were fragmented in fragmentation buffer, then short fragments were subjected to reverse transcription and second-strand synthesis. The cDNA fragments were treated with standard library generation steps; end-repair, A-tailing, and adapter ligation. After purification with DNA clean beads, we enriched adapter-ligated fragments using 14 PCR cycles and subjected to the following denaturation and single-strand circularization process to generate a single-stranded circular DNA library. These libraries were then used to generate DNA nanoballs (DNBs) by rolling circle replication (RCR). The resulting DNBs were then loaded into the patterned nanoarrays, and sequencing reaction was performed in a DNBSEQ-G400 sequencer with a pair-end read length of 100 bp.

### RNA-Seq Data Analysis and Differential Expression Analysis

The quality of raw sequencing reads was checked with FastQC (Babraham Bioinformatics) before and after sequence trimming. For a comparison of the qualities of all RNA-Seq libraries, MultiQC [25] software was used to merge the results of FastQC. Raw reads were filtered using fastp v0.23.0 [26] to remove adaptor contamination, ambiguous (N>5) bases, low quality reads (Phred score, *Q*<20), and fragments <30 nt. All other options used the default values. Summary statistics for RNA-Seq reads were computed using seqkit v2.0.0 (Shen et al., 2016). Filtered reads were mapped to human reference genome (GRCh38.p13, Ensembl Release 106) using Hisat2 v2.2.1 [27]. The alignment statistics were obtained with Sambamba v0.8.0 [28]. Count matrices and gene-level assignment were generated using featureCounts from Subread package v2.0.0 [29] with annotation version GRCh38.106 (Ensembl “.gtf”). Differential gene expression between groups was performed on raw counts using DESeq2 v1.34.0 [30] after variance-stabilizing transform (vst) normalization. Genes were considered as significantly differentially expressed if the adjusted *P*Lvalue (Benjamini– Hochberg (BH) multiple test correction method) was less than 0.001 and log_2_FC>1.0. Hierarchical clustering and principal component analysis (PCA) were performed using DEBrowser v1.20.0 [31] to evaluate the correlation between control and disease samples. Volcano plots of most differentially expressed genes (DEGs) among comparison groups were generated using EnhancedVolcano v1.12.0 [32]. The top 34 up-regulated and down-regulated protein-coding genes (sorted by adjusted *p*-value in increasing order) for each comparison were selected for heatmap generation using the online tool ClustVis[33]. The multiple plots generated from each condition were combined one overall graph using ggarrange() function available in the ggpubr R package (https://rpkgs.datanovia.com/ggpubr/).

### Functional Annotation and Enrichment Analysis

Gene ontology (GO) and the Kyoto Encyclopedia of Genes and Genomes (KEGG) pathway enrichment analysis of DEG among groups of control, no pneumonia and severe pneumonia was performed with Metascape using *Homo sapiens* (Ensembl Release 104) as background for enrichment[34]. The KEGG pathway enrichment analysis of DElncRNAs among groups of control, no pneumonia and severe pneumonia was performed with NCPATH using *Homo sapiens* (Ensembl Release 104) as background for enrichment[35]. Significance of enrichment analysis was estimated by Benjamini–Hochberg false discovery rate (FDR) <0.05 correction. Protein-protein interaction (PIP) network analysis was performed sing STRINGdb v12.0 [36]. Normalized expression data were used for discovery co-expressed modules, gene-diseases interaction, gene interactions with transcription factors and their targets, immune cell signatures using Metascape[34] and Enrichr [37, 38]. A variance filter value of 0.01 was used to ensure the highest level of statistical stringency and Pearson’s correlation method was selected for identification of the gene modules.

### Identification of 52 Genes Associated with Idiopathic Pulmonary Fibrosis Risk in The PBMCs’ Transcriptome

In our PBMC transcriptome data, we compared up and down profiles based on the expression levels of 7 increased genes (PLBD1, TPST1, MCEMP1, IL1R2, HP, FLT3, S100A12) and 45 decreased genes (LCK, CAMK2D, NUP43, SLAMF7, LRRC39, ICOS, CD47, LBH, SH2D1A, CNOT6L, METTL8, ETS1, P2RY10, TRAT1, BTN3A1, LARP4, TC2N, GPR183, MORC4, STAT4, LPAR6, CPED1, DOCK10, ARHGAP5, HLA-DPA1, BIRC3, GPR174, CD28, UTRN, CD2, HLA-DPB1, ARL4C, BTN3A3, CXCR6, DYNC2LI1, BTN3A2, ITK, CD96, GBP4, S1PR1, NAP1L2, KLF12, IL7R, SNHG1, C2orf27A) from a gene signature previously found to be predictive of IPF poor prognosis and COVID-19 outcome [39–41]. The genes (sorted by adjusted p-value in increasing order) for each comparison were selected for heatmap generation using the online tool ClustVis[33].

### Statistical Analysis

Data were checked for normality and continuous variables were compared using one-way variance analysis, ANOVA/Dunnett’s multiple comparison tests, or Kruskal-Wallis/Dunn’s multiple comparison tests in the context of the RNAseq count data-based on gene expression analysis of the hub genes. The findings are presented as medianL±Linterquartile (IQ) ranges or meanL±LSD. P values were considered significant if they were less than 0.05. PRISM version 8 (GraphPad Software Inc, San Diego, CA, United States) was used for the statistical analysis.

## 3. Results

### 3.1. Identification of DEGs and DElncRNAs among Healthy Control, No Pneumonia and Severe Pneumonia Groups

The success of the transcriptome sequencing reaction depends on the quality and quantity of the RNA isolated. Of the 40 PBMC-RNA samples sent for transcriptomic analysis, 36 samples with high quality RNA were analysed, except for three patients with severe pneumonia and one patient without pneumonia. The RNA samples of the other 2 patients without pneumonia were not included in the analysis because it was determined that they could not reach the reading quality with sufficient number and accuracy for bioinformatic analysis. After the bioinformatic analysis of the raw data obtained after the sequence analysis, a total of 34 RNA samples from the control group (N=13), the post-Covid 19 group without pneumonia (N=10) and the post-Covid-19 group with severe pneumonia (N=11) were included in the analysis because the reading quality was of sufficient quality (**Figure 1a**). The relative status of gene expression changes in all three groups is shown by PCA analysis using the transcriptomic gene expression profile from PBMCs from control (n=13), no pneumonia (n=10), and severe pneumonia (n=11) individuals. As a result, we observed that the control samples grouped apart from the other two groups and that the gene expression variations in people with no and severe pneumonia differed more than those in the control group (**Figure 1b**). Moreover, a cluster heat map was created from 34 upregulated and 34 downregulated genes to show differential gene expression in each group comparisons (**Figure 1c**). In the comparison between healthy control (C) and no pneumonia group (NP), we identified 4843 DEGs including 3004 upregulated (up) genes and 1839 downregulated (down) genes, and 1056 DElncRNAs including 694 up and 392 down lncRNAs (**Figure 2a-c**; **Figure 6a-c**). We identified 1651 DEGs including 1566 up and 85 down genes, and 577 DElncRNAs including 493 up and 84 down lncRNAs between healthy control and severe pneumonia group (C versus SP). In NP vs SP comparison, we identified 954 DEGs including 79 up and 875 down genes, and 148 DElncRNAs including 5 up and 143 down lncRNAs (**Figure 2a-c**; **Figure 6a-c**). We then overlapped DEGs from all comparisons. Totally, we identified 291 DEGs and 70 DElncRNAs. Additionally, Upregulated and Downregulated DEGs and DElncRNAs were showed in Venn diagrams. Next, we tried to identify the function of common DEGs and DElncRNAs involved during the pneumonia triggered by Long Covid-19 (**Figure 2a-c**; **Figure 6a-c**).

**Figure 1.**
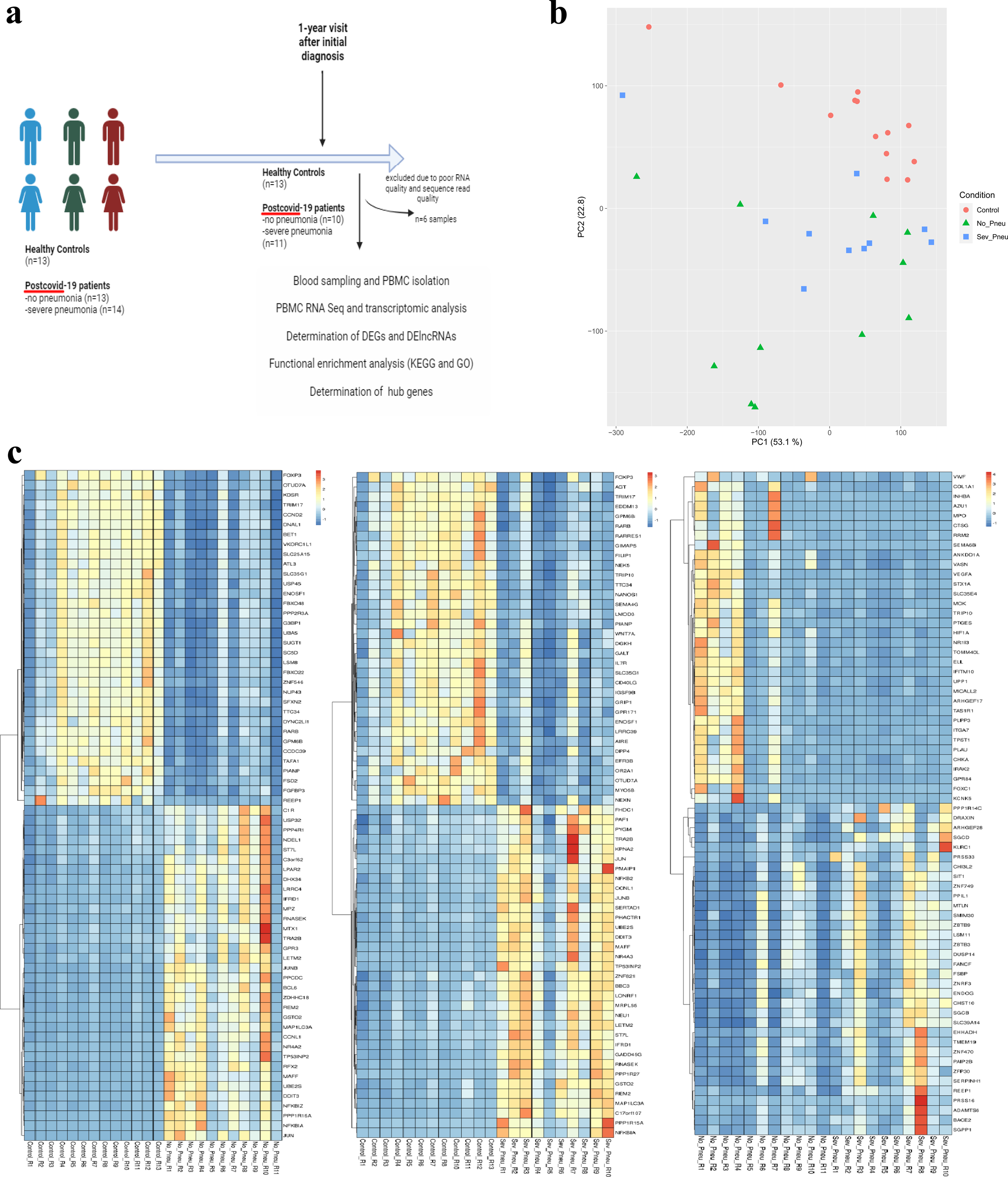
**(a)** A graphical abstract of the study. **(b)** Principal component analysis is shown. Each point represents a sample. Colouring indicates groups. Red circles represent the control samples, green triangles represent the patient samples with COVID-19 disease from a year ago and no pneumonia in the follow-up radiological data, and blue squares represent the samples of patient with COVID-19 disease from a year ago and severe pneumonia confirmed by radiological data in the follow-up period. (**c)** Heatmap of 34 upregulated and 34 downregulated DEGs in all group comparisons.

**Figure 2.**
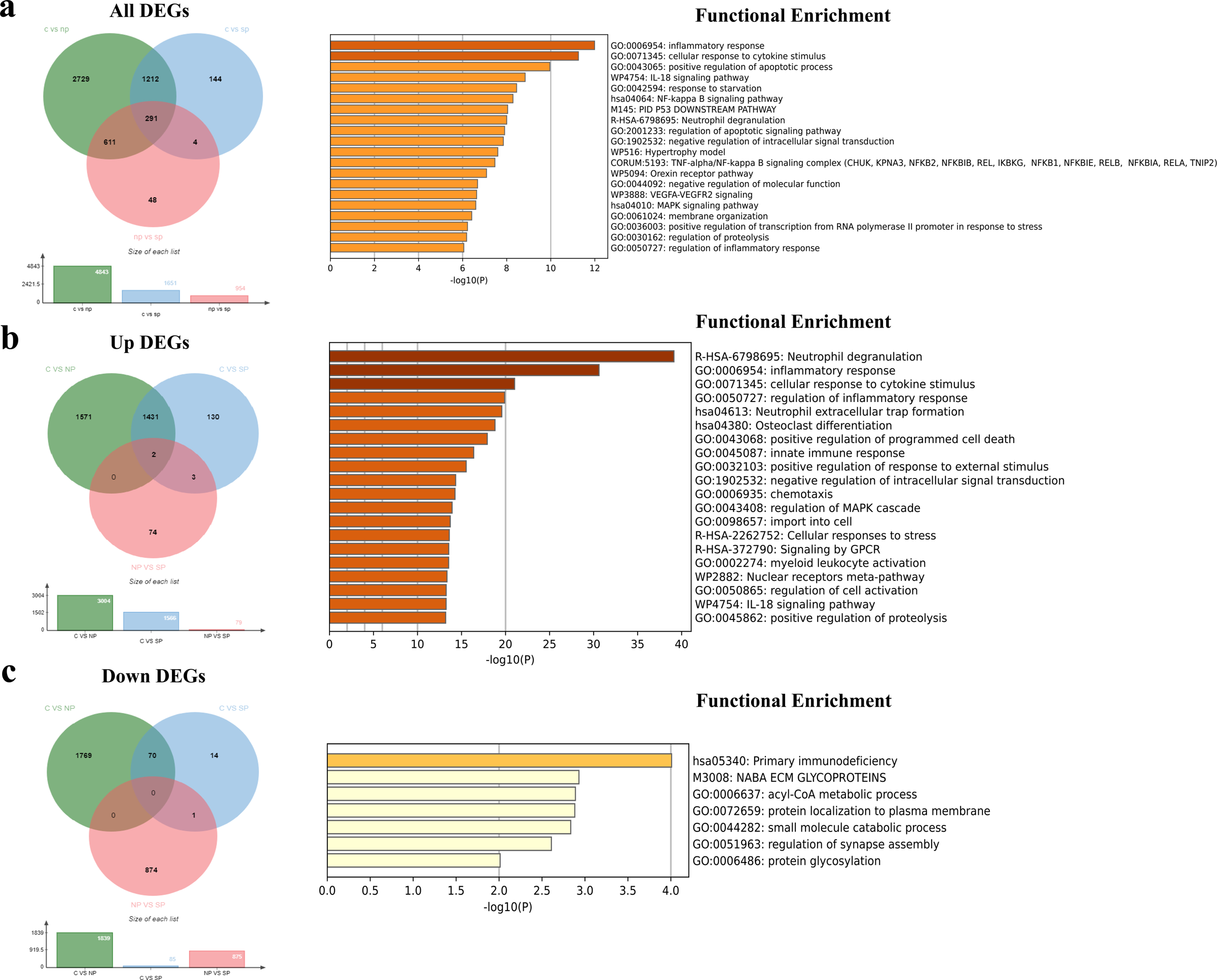
**(a)** All differentially expressed genes (DEGs) comparison among “C vs NP”, “C vs SP” and “NP vs SP”, and common 291 DEGs. Functional enrichment analysis (Kyoto Encyclopedia of Genes and Genomes (KEGG), Gene Ontology (GO), WikiPathways (WP), and the comprehensive resource of mammalian protein complexes (CORUM)) of common 291 DEGs. **(b)** Upregulated DEGs among all comparisons, and 1436 extended common upregulated genes. Functional enrichment analysis of common 1436 up-DEGs. **(c)** Downregulated DEGs among all comparisons, and 71 extended common downregulated genes. Functional enrichment analysis of common 71 down-DEGs. The green circle in Venn diagram represents DEGs in “C vs NP” comparison, and blue circle represents DEGs in “C vs NP” comparison, and red circle represents DEGs in “NP vs SP” comparison.

### 3.2. Functional Enrichment Analysis of all DEGs

To understand the function and pathways of all DEGs, enrichment analysis was performed. This analysis revealed that upregulated DEGs in “C vs NP” comparison was involved in osteoclast differentiation (KEGG) and positive regulation of inflammatory process (GO:0050727). The DEGs downregulated in the comparison, however, were associated with herpes simplex virus 1 infection (KEGG) and the positive regulation of natural killer cell mediated immunity (GO:0002717). The upregulated DEGs were mainly located in tertiary granules, and the main molecular function of these genes was found as protein serin/threonine kinase activity. The downregulated DEGs were mainly located in sarcoglycan and dystroglycan complexes, and the main molecular function of these down genes was found to bind to DNA via RNA polymerase II transcription regulatory region sequence **(Figure S1a)**. Upregulated DEGs in “C vs SP” comparison was involved in osteoclast differentiation (KEGG) and positive regulation of inflammatory process (GO:0050727). The DEGs downregulated in the comparison, however, were associated with primary immunodeficiency (KEGG) and alcohol catabolic process (GO:0046164). The upregulated DEGs were mainly located in secretory granule membrane, and the main molecular function of these genes was found as cytokine receptor activity and G protein coupled receptor activity. The downregulated DEGs were mainly located in keratin and intermediate filaments, and the main molecular function of these down genes was found as oxidoreductase activity acting on NAD or NADP as an acceptor **(Figure S1b)**.

Upregulated DEGs in “NP vs SP” comparison was involved in herpes simplex virus 1 infection, viral myocarditis, arrhythmogenic right ventricular cardiomyopathy, sulphur metabolism, hypertrophic cardiomyopathy, and TGF-beta signaling pathway (KEGG) in addition to the regulation of lymphocyte activation (GO:0051249), and some cardiac tissue morphogenesis processes (such as GO:0048738, GO:0061384, GO:0051146). The DEGs downregulated in the comparison, however, were associated in the transcriptional misregulation in cancer and TNF signaling (KEGG), and the positive regulation of transcription by RNA polymerase II (GO:0045944). The upregulated DEGs were mainly located in sarcoglycan and dystroglycan complexes, and the main molecular function of these genes was found as serine-type endopeptidase activity. The downregulated DEGs were mainly located in specific and azurophil granules, and the main molecular function of these genes was found as protein serine/threonine kinase activity. **(Figure S1c)**.

### 3.3. Functional Enrichment Analysis of Common DEGs and DElncRNAs

Enrichment analysis was conducted to gain a deeper understanding of the function and pathways of common DEGs. The results indicated that 291 common DEGs across all DEGs were primarily engaged in inflammatory response processes, including TNF-alpha, NF-κB, and MAPK signalling pathways (**Figure 2a**). Moreover, the extended common 1436 DEGs in upregulated DEGs were involved in neutrophil degranulation, neutrophil extracellular trap formation, and osteoclast differentiation (KEGG), and cellular inflammatory response (GO: 0006954 and GO:0071345) (**Figure 2b**). Furthermore, the extended common 71 DEGs in downregulated DEGs were mainly involved in primary immunodeficiency (KEGG), and some metabolic processes such as acyl-CoA process (GO: 0006637), small molecule catabolic process (GO: 0044282), and protein glycolisation (GO: 0006486) (**Figure 2c**). The results indicated that 70 common lncRNAs across all DElncRNAs were primarily engaged in MAPK and Rap1 signaling pathways (KEGG). Moreover, the extended common 457 lncRNAs in upregulated DElncRNAs were involved in MAPK signaling pathway, focal adhesion, cell cycle, and insulin resistance (KEGG). Furthermore, the extended common 79 lncRNAs in downregulated DElncRNAs were mainly involved in thermogenesis and mTOR signaling pathway (KEGG). Finally, we revealed that 23 common DEGs and 70 DElncRNAs were enriched in MAPK, Rap1, and AMPK signaling pathways (KEGG) **(Table S1)**.

### 3.3. Identification of Hub Genes via Protein–Protein Interaction (PPI) Network

The PPI network analysis revealed an association of DEGs among healthy control, no pneumonia and severe pneumonia groups. We identified 291 common DEGs consisted of 291 nodes with 142 edges (**Figure 3a**). Thirty-six genes were determined from the 291 common genes using Metascape online software. Among 36 genes, the highest scoring (Interactions≥3) in 23 central genes, namely ICAM1, TUBB4B, MARCKS, NFKB2, NFKBIA, NFKBIE, HDAC5, ATF3, DDIT3, F3, PRKCD, IL1R1, AREG, CSF1, IL1R2, TOM1, RAB11FIP1, FSCN1, ULK1, RELB, NFKBIB, FOSL1, and JUND were identified from the PPI network in combination with the Metascape (**Table 2**). When we analysed these 23 genes, we obtained a hub gene network with a total of 14 genes in 2 modules (**Figure 3c**), and then, we revealed that these genes were specifically involved in the TNF-alpha/NF-κB signaling complex, osteoclast differentiation, and cytokine signaling in the immune system.

**Figure 3.**
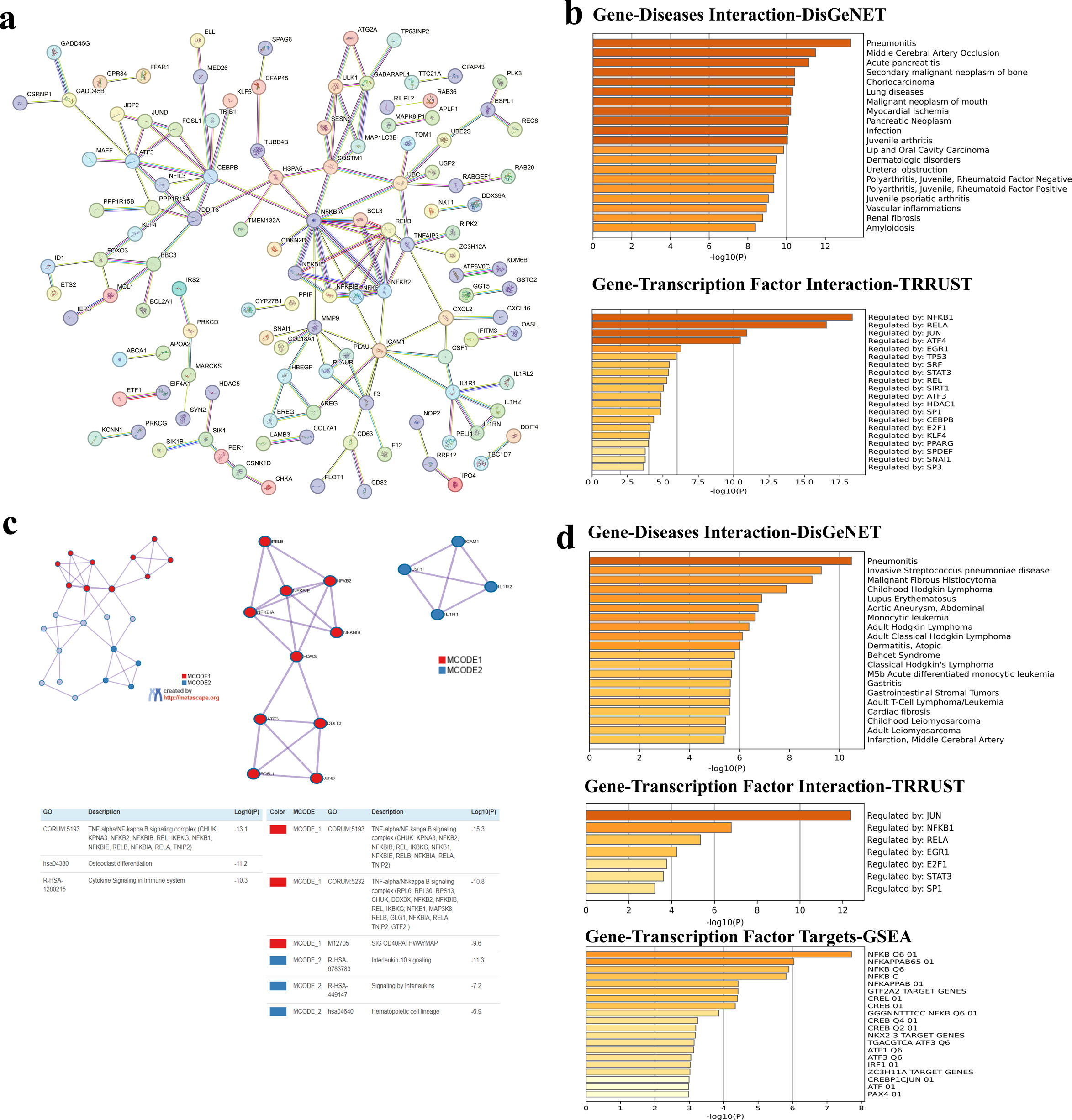
**(a)** Protein-protein interaction analysis of 291 common genes determined as a result of comparison of DEGs in all comparable groups. **(b)** Gene-Diseases (DisGeNET) and Gene Transcription Factor interactions (TRRUST) of these genes. **(c)** PPI analysis of hub genes from 291 genes. **(d)** Gene-Diseases (DisGeNET), Gene Transcription Factor interactions (TRRUST), and gene-transcription factor targets (GSEA) of these hub genes. The graph shows the diseases, transcription factors and their targets with which DEGs are highly correlated in these intergroup comparisons. The vertical axis shows the disease names obtained from the DisGenet database, and the horizontal axis shows the −log10 (P) statistical significance level.

**Table 2.** Following the comparative analysis of all up-regulated and down-regulated genes across the control, no pneumonia, and severe pneumonia groups, 36 hub genes were identified from a total of 291 common DEGs. Subsequently, a selection of 23 central genes, each interacting with three or more other genes, was made, along with an investigation into the genes, which they interact with.

| **Gene Symbol** | **Number of genes interacted** | **Names of genes interacted** |
| --- | --- | --- |
| **ICAM1** | 6 | AREG, CSF1, IL1R1, IL1R2, PRKCD, TUBB4B |
| **MARCKS** | 6 | F3, FSCN1, PRKCD, RAB11FIP1, TUBB4B, ULK1 |
| **TUBB4B** | 6 | F3, GABARAPL1, ICAM1, MARCKS, SNAI1, ULK1 |
| **HDAC5** | 5 | ATF3, DDIT3, NFKB2, NFKBIA, NFKBIE |
| **NFKBIE** | 5 | HDAC5, NFKB2, NFKBIA, NFKBIB, RELB |
| **NFKB2** | 5 | HDAC5, NFKBIA, NFKBIB, NFKBIE, RELB |
| **NFKBIA** | 5 | HDAC5, NFKBIB, NFKBIB2, NFKBIE, RELB |
| **ATF3** | 4 | DDIT3, FOSL1, HDAC5, JUND |
| **F3** | 4 | ICAM1, MARCKS, PRKCD, TUBB4B |
| **DDIT3** | 4 | ATF3, FOSL1, HDAC5, JUND |
| **IL1R1** | 4 | CSF1, ICAM1, IL1R2, TOM1 |
| **PRKCD** | 4 | F3, FSCN1, ICAM1, MARCKS |
| **AREG** | 3 | ERG, HBEGF, ICAM1 |
| **CSF1** | 3 | ICAM1, IL1R1, IL1R2 |
| **FOSL1** | 3 | ATF3, DDIT3, JUND |
| **FSCN1** | 3 | MARCKS, MCL1, PRKCD |
| **IL1R2** | 3 | CSF1, ICAM1, IL1R1 |
| **JUND** | 3 | ATF3, DDIT3, FOSL1 |
| **NFKBIB** | 3 | NFKB2, NFKBIA, NFKBIE |
| **RAB11FIP1** | 3 | IFITM3, MARCKS, TOM1 |
| **RELB** | 3 | NFKB2, NFKBIA, NFKBIE |
| **TOM1** | 3 | IL1R1, IFITM3, RAB11FIP1 |
| **ULK1** | 3 | GABARAPL1, MARCKS, TUBB4B |
| **ERG** | 2 | AREG HBEGF |
| **GABARAPL1** | 2 | TUBB4B, ULK1 |
| **HBEGF** | 2 | AREG, ERG |
| **IFITM3** | 2 | RAB11FIP1, TOM1 |
| **MCL1** | 2 | FSCN1, USP2 |
| **SNAI1** | 2 | TUBB4B, USP2 |
| **USP2** | 2 | MCL1, SNAI1 |
| **GADD45B** | 2 | GADD45G, MAP2K3 |
| **GADD45G** | 2 | GADD45B, MAP2K3 |
| **MAP2K3** | 2 | GADD45B, GADD45G |
| **CSNK1D** | 2 | HSPA5, NOP2 |
| **HSPA5** | 2 | CSNK1D, NOP2 |
| **NOP2** | 2 | CSNK1D, HSPA5 |

### 3.3. Identification of Gene-diseases, Gene-transcription factors, and Gene-transcription factor Targets Interactions

Gene-Diseases interaction via DisGeNET interestingly showed that the 291 common genes were related to pneumonitis, middle cerebral artery occlusion, acute pancreatitis, secondary malignant neoplasm of bone, choriocarcinoma, lung diseases, malignant neoplasm of mouth, myocardial ischemia, pancreatic neoplasm, infection and juvenile arthritis. We determined that these genes consisted of genes expressed especially under the control of NFKB1, RELA, JUN and ATF4 transcription factors (**Figure 3b**). Our remarkable discovery was that the 14 hub genes we identified were primarily related to pneumonitis according to DisGeNET analysis. In our analysis, we saw that these genes, especially those carrying NFKB Q6, one of the transcription factor targets, were under the control of JUN (**Figure 3d**).

### 3.4. Identification of Immune Cell Type Signatures of the Common and Central DEGs

To determine immune cell type signatures in the extended common 1436 upregulated, the extended common 71 downregulated, 291 common, and 23 central DEGs, our Metascape analysis showed that the 1436 DEGs mainly involved in neutrophil cells and monocytes (**Figure 4a**). Downregulated common 71 DEGs mainly involved in CD4 memory effector T cells (**Figure 4b**). The common 291 DEGs involved in monocytes, dendritic cells, basophils, and neutrophils (**Figure 4c**). Interestingly, 23 central genes mainly involved in CCL19 and CCL21 positive cells, monocytes, basophils, and dendritic cells (**Figure 4d**).

**Figure 4.**
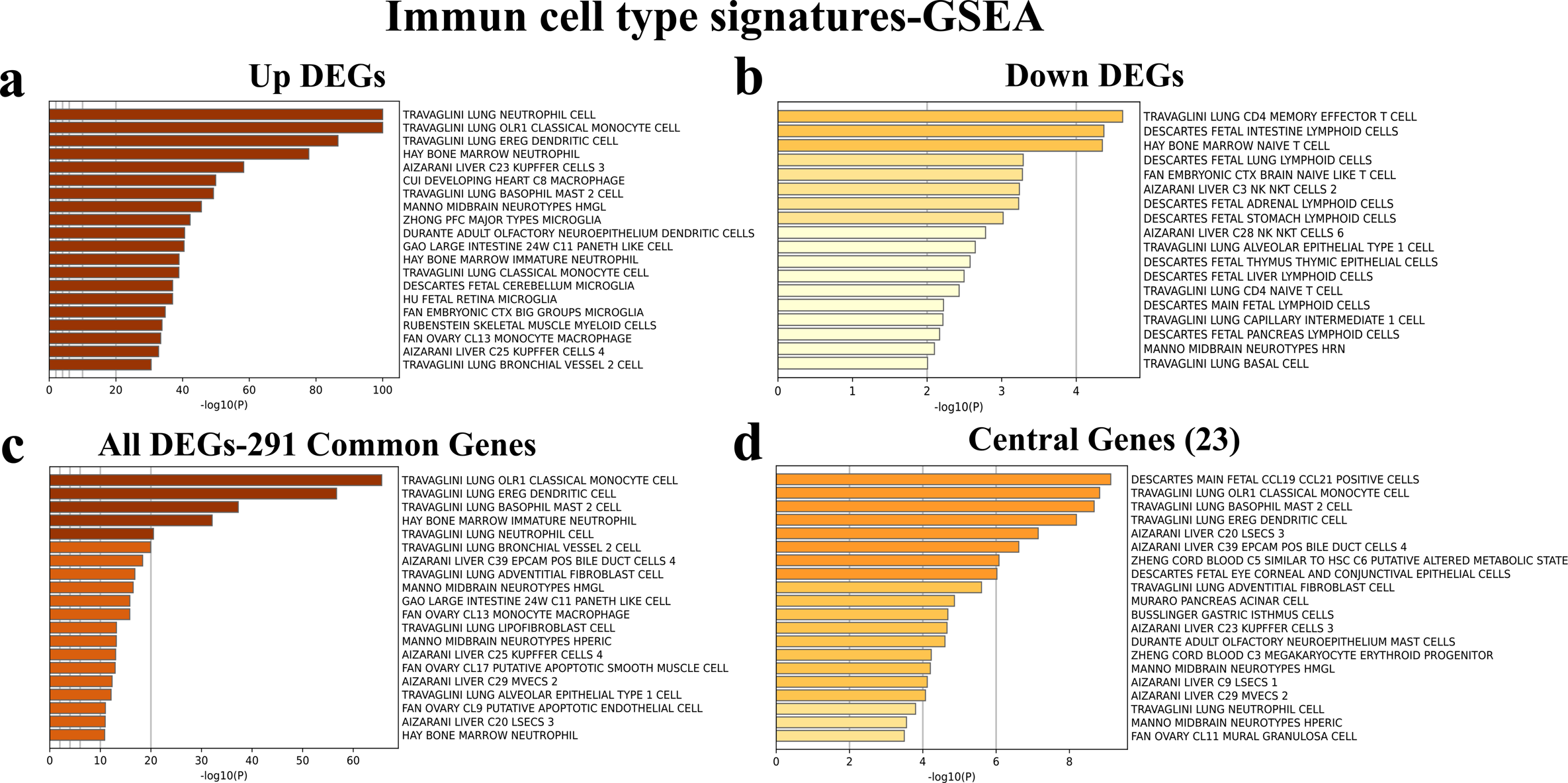
Immune cell signatures of **(a)** the extended common upregulated DEGs, **(b)** the extended common downregulated DEGs, **(c)** the 291 common DEGs, and **(d)** the 14 hub genes. The graph shows the immune cell signatures with which DEGs are highly correlated in these intergroup comparisons. The vertical axis shows the immune cell names obtained from the databases, and the horizontal axis shows the −log10 (P) statistical significance level.

### 3.5. Gene Expression Analysis Based on RNA-seq Count Data of 14 Hub Genes

We performed a gene expression analysis based on statistical analysis of the normalized counts of the hub genes and GAPDH of each patient and control. According to these results, all genes were significantly upregulated in no pneumonia groups compared to healthy control individuals. Also, the other genes except *FOSL1* and *CSF1* were significantly upregulated in severe pneumonia group compared to the control groups. *FOSL1* (p<0.05) and *CSF1*(p<0.01) genes were downregulated in severe pneumonia group compared to no pneumonia group. In other genes, no significance was found between no pneumonia and severe pneumonia (**Figure 5**).

**Figure 5.**
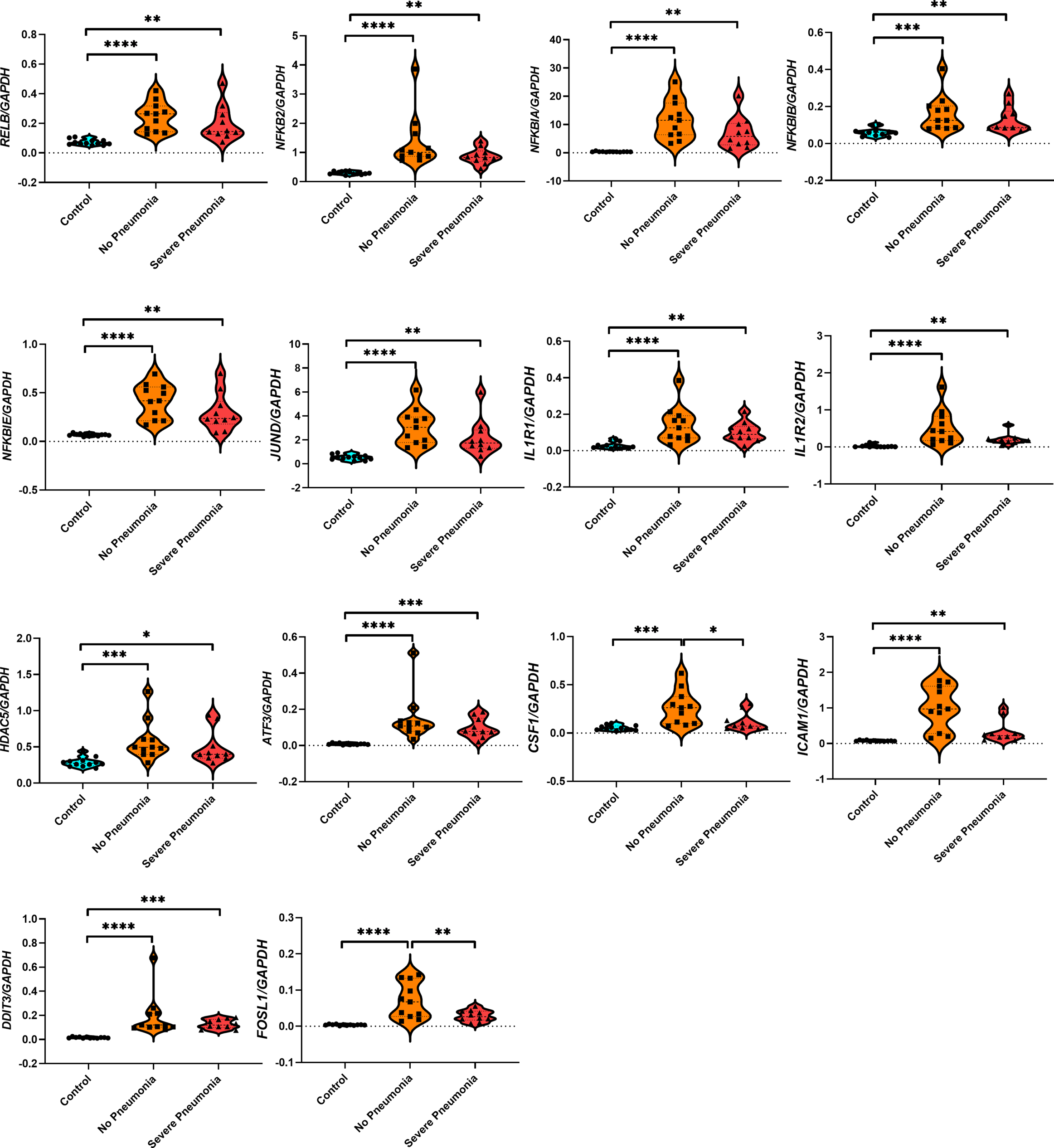
Gene expression analysis based on RNA-seq count data of 14 hub genes. All target genes’ counts were normalized the counts of the individual GAPDH. Blue and circles shows control individuals, Orange and squares shows the individuals with no pneumonia, and red and triangles shows the individuals with severe pneumonia. Statistical significance was shown as *p<0.05, **p<0.01, ***p<0.001, and ****p<0.0001.

### 3.6. Investigation of Idiopathic Pulmonary Fibrosis related 52 Gene Signatures in PBMC Transcriptome of Patients with Non-Pneumonia and Severe Pneumonia

The transcriptome signals of the PBMCs from individual patients in the "NP " and "SP" groups were analysed using a predefined set of 52 PBMC signature genes and visualised in a heatmap to determine whether these groups exhibited IPF signals. Our results shown that the expression levels of the genes PLBD1, TPST1, MCEMP1, IL1R2, HP, FLT3, and S100A12, which were low in the control group, significantly increased in patients, who had COVID-19 but did not develop pneumonia (’NP’). In patients with severe pneumonia due to COVID-19 (’SP’), an increase was observed in PLBD1, S100A12, and MCEMP1 genes, while no significant changes were noted in the other four genes **(Figure S1d)**. Also, we demonstrated that the expression levels of the genes LCK, CAMK2D, NUP43, SLAMF7, LRRC39, ICOS, CD47, LBH, SH2D1A, CNOT6L, METTL8, ETS1, P2RY10, TRAT1, BTN3A1, LARP4, TC2N, GPR183, MORC4, STAT4, LPAR6, CPED1, DOCK10, ARHGAP5, HLA-DPA1, BIRC3, GPR174, CD28, UTRN, CD2, HLA-DPB1, ARL4C, BTN3A3, CXCR6, DYNC2LI1, BTN3A2, ITK, CD96, GBP4, S1PR1, NAP1L2, KLF12, IL7R, SNHG1, and C2orf27A, which were high in the control group, significantly decreased in both NP and SP groups (Figure S1d).

## 4. Discussion

Subsequent to the propagation of COVID-19, the observation of protracted health complications among individuals recuperating from the acute phase of the disease has engendered novel and complex challenges for the scientific community and healthcare professionals. Termed as "Long-COVID," which is characterized by persistent symptoms that pervasively affect various organ systems, culminating in a substantial diminution of patients’ quality of life. These symptoms encompass fatigue, dyspnoea, pneumonia, pulmonary fibrosis, and pervasive inflammatory responses[8]. The multi-omics analyses of whole blood and PBMC samples have been instrumental in elucidating the cellular signals underpinning the disease’s mechanisms and symptoms, contributing significantly to parallel research endeavours [42] [43].

In our study, we conducted a comprehensive analysis of RNA sequences from PBMCs to interrogate the differential gene and lncRNA expression profiles amongst healthy controls, Lost Covid-19 patients without pneumonia (NP), and Long-COVID-19 patients with severe pneumonia (SP). The investigation’s main finding indicates that, regardless of pneumonia manifestation, the TNF-alpha/NFκB signalling complex and MAPK signalling pathways regulate the positive control of inflammatory processes one year after COVID-19. This modulation, together with neutrophil-associated signatures, could be the underlying cause of the reported problems in patients. In one of the pioneering studies, proteomic analysis was conducted with blood plasma collected for two 6-month visits, and multiplex gene expression analysis was performed with RNA obtained from nasal epithelial cells. This study showed the presence of elevated systemic inflammatory signals even 3-6 months post-disease, and reported that the radiological and functional alterations observed in these individuals did not revert to normal within a 12-month period [44]. These findings corroborate the alterations observed in our studies. However, our investigation is distinguished as the inaugural transcriptomic analysis conducted with PBMCs isolated from blood samples obtained at the end of one year, from randomly selected long-Covid-19 patients with no pneumonia, severe pneumonia, and healthy controls, matched for age and gender. The PCA analysis and cluster heat maps provided a clear visual representation of the variations in gene expression, with the control samples distinctly separating from the patient groups. This separation is indicative of a fundamental shift in gene expression dynamics in Long-COVID-19, which could be instrumental in understanding the lingering effects of the disease and potential avenues for therapeutic interventions. The identification of DEGs revealed distinct molecular signatures between the groups. Based on group comparisons of healthy control vs no pneumonia, healthy control vs severe pneumonia, and no pneumonia vs severe pneumonia, DEGs that were upregulated in both no pneumonia and severe pneumonia patients compared to control individuals were enriched in osteoclast differentiation, HIF1α, IL-17 and TNF signaling pathways, while downregulated genes were found to be enriched in Herpes simplex virus 1 infection in the "C vs NP" comparison and primary immunodeficiency in the "C vs SP" comparison. This situation suggests the long-lasting effect observed following excessively increased inflammatory signals after virus infection. According to the no pneumonia group, it was determined that the upregulated genes in patients with severe pneumonia observed in their radiology and clinics were mainly clustered in herpes simplex virus 1 infection and cardiac changes secondary to the virus, while the downregulated genes were especially found to be associated with transcriptional mis-regulation in cancer and decrease in TNF-α signaling. The "NP vs SP" comparison revealed a unique set of DEGs involved in cardiac-related processes and viral infections, highlighting the potential cardiovascular risks and lingering susceptibility to viral infections in Long-COVID-19 patients.

Previous investigations have elucidated that TNF-α accelerates osteoclast differentiation independently through the receptor activator of NF-κB ligand (RANKL), RANK, and TRAF6, while IL-17A facilitates the same process via RNAKL-JNK1 signaling pathway[45, 46] [47]. Studies have reported that the differentiation of osteoclasts is intricately coordinated through the binding of macrophage colony-stimulating factor (M-CSF) and RANKL to their respective receptors on the surface of osteoclast precursor cells. It has been documented that M-CSF plays a pivotal role in the proliferation of osteoclasts, whereas RANKL is instrumental in their differentiation[48].

In a murine model study, silica-induced pulmonary fibrosis was observed to stimulate osteoclast-like differentiation, leading to the recruitment of monocytes to the tissue. This process of osteoclast-like differentiation of alveolar macrophages is mediated by the osteoclastogenic cytokine RANKL, which is released from pulmonary lymphocytes and type II alveolar epithelial cells [49]. Collectively, these studies suggest the potential involvement of this mechanism in the persistent effects observed post-viral infection.

The identification of common DEGs and DElncRNAs across all comparisons, as well as the elucidation of hub genes, provided a holistic view of the pivotal molecular players in Long-Covid-19 conditions. In present study, the enrichment of these common DEGs in inflammatory response processes and signaling pathways, such as TNF-α, NF-κB, and MAPK, underscores the persistent inflammatory state and potential for chronic complications in Long-Covid-19 patients. This augmentation of signatures was already expressed in an editorial [50]. The PPI network analysis and identification of hub genes further highlighted the interconnectedness of these molecular players, with genes such as *RELB, NFKB2, NFKBIA, NFKBIB, NFKBIE, HDAC5, ATF3, DDIT3, FOSL1, JUND, CSF1, ICAM1, IL1R1* and *IL1R2* emerging as central nodes in the network. These hub genes are known to play crucial roles in immune responses and inflammation, reinforcing the idea that Covid-19 conditions are characterized by prolonged immune activation and potential dysregulation. In a multi-omics analysis performed on the pulmonary tissues of COVID-19 patients [51], the study elucidated that the complications observed in the acute phase of the disease were underpinned by augmented mechanisms of senescence, inflammation, apoptosis, coagulation, and fibrosis. Furthermore, the genes and proteins playing a role in these mechanisms, as well as their associated signaling networks, were elucidated. Intriguingly, within these networks, certain molecular entities (RELB, DDIT3, and FOSL1) were identified as hub genes with increased expression in our patient samples during the 1-year Long-COVID-19 follow-up period. These genes are thought to play a role in ongoing pneumonia and are believed to bridge the networks of apoptosis, inflammation, and fibrosis [51]. The continued upregulation of these genes, initially detected in the acute phase, throughout the 1-year recovery period Long-COVID-19 is noteworthy. This phenomenon could potentially lead to the emergence of pneumonia complications and other inflammation-related outcomes.

Interestingly, our gene-diseases interaction analysis provided intriguing links between the common DEGs and various diseases, including especially pneumonitis, other lung-related conditions, and arthritis. Transcriptomic study of the acute phase PBMCs of COVID-19 patients revealed a number of immune-related illnesses, such as pneumonia, arthritis, and septicemia, in earlier research[43]. As far as is known, the lung is the organ primarily affected by SARS-CoV-2 infection. SARS-CoV-2 spreads in the lower respiratory tract of the severe patients, resulting in hypoxemia, severe pneumonia, and acute respiratory distress syndrome[52]. When single-cell RNA-seq analyses were conducted on pathological samples taken from the lungs of COVID-19 patients during the acute phase, they found strong inflammation and cellular markers associated with apoptosis-dependent fibrotic alterations. They hypothesised that these signals could be connected to inflammation and fibrotic alterations brought on by acute pneumonia [51].

The immune cell type signature analysis revealed distinct profiles between the upregulated and downregulated DEGs, with neutrophils and monocytes dominating the upregulated DEGs and CD4 memory effector T cells being prominent among the downregulated DEGs. Transcriptomic analyses performed on the PBMCs of COVID-19 patients in the acute phase have reported an enrichment of upregulated genes in bone marrow and blood CD33^+^ myeloid cells as well as CD14^+^ monocytes, while downregulated genes were observed to be enriched in CD4 and CD8 T cells, as we have also noted [43]. This shift in immune cell type signatures could have implications for the patient’s immune response and susceptibility to secondary infections in Long-COVID-19. The gene expression analysis of the 14 hub genes provided a granular view of their expression patterns across the groups, with significant upregulation observed in the no pneumonia group compared to healthy controls. This upregulation, coupled with the distinct expression patterns in the severe pneumonia group, suggests a complex interplay of molecular factors contributing to the varied clinical outcomes in Long-COVID-19 patients. The investigation of idiopathic pulmonary fibrosis (IPF) related gene signatures revealed a subset of genes with altered expression in the no pneumonia and severe pneumonia groups, providing a potential link between COVID-19 and the risk of developing IPF-like conditions. In our investigation, all seven genes exhibited increased expression in COVID-19 patients without pneumonia, while only certain genes like PLBD1, S100A12, and MCEMP1 showed increased expression in severe pneumonia cases. These seven genes’ upregulation correlates with severe IPF and COVID-19 outcomes, as evidenced by prior studies [39–41]. Additionally, all 45 genes displayed decreased expression across both patient groups and have been previously associated with severe IPF and COVID-19 risks. The NP group presented transcriptomic signatures similar to IPF [41]. We identified these high-risk genes predominantly in CD14^+^ monocytes, dendritic cells, and neutrophils, highlighting their pivotal role in mediating high-risk gene-associated responses.

## 5. Conclusion

In summary, our study provides a comprehensive transcriptomic analysis of PBMCs in post-Covid-19 conditions, uncovering a complex landscape of DEGs, DElncRNAs, and hub genes associated with various clinical outcomes. The persistent inflammatory state, alterations in immune cell type signatures, and potential links to lung-related conditions and IPF highlight the need for ongoing research and monitoring of post-Covid-19 patients. Future studies should focus on validating these findings in larger cohorts, exploring the therapeutic implications of these molecular alterations, and investigating the long-term health impacts of Covid-19 to enhance our understanding and improve the care of affected individuals.

## Author contributions

OK, HB, VE, SAB, NK, DM, OBT, PDC, FF and OI designed the study; SAB, SCK, OBT, PDC, FF, NK, DM, SK, OK, YT, EA, MK, PAY, PPD, OK, IB, HB, IH, NK, GS, CC, HKO, HSO, BE, MH, AS, ENT, OO, TUC, IKO, VAO, FB, OA, ME, PYG, ATE, MMT, OI and HB collected the data; OK, VE, SCK, HB analysed the data; OK, HR, NK, DM, SKK, SE, GTA, GE and YT searched the literature and OK wrote the manuscript; SAB, OBT, FF, PDC, NK and HB edited and revised manuscript according to journal’s instructions; All authors edited and controlled the final version of the manuscript. All the authors approved the final version of the manuscript.

## Conflict of interest statement

The authors have no conflict of interest to declare.

## Funding

This study was funded by the Research Support Fund of the Turkish Thoracic Society (TTD).

## Data Availability

All data produced in the present study are available upon reasonable request to the authors. FASTQ sequences of the PBMC samples have been deposited in NCBI Short Read Archive (SRA) under BioProject PRJNA895325.

## Acknowledgments

We greatly appreciate the patients and healthy volunteers involved in the study. Also, we would like to thank all of the TURCOVID and POST-COVID collaborators, who cooperated for this study. We would like to thank the Turkish Thoracic Society for the funding and support. Furthermore, the authors acknowledge the use of the services and facilities of the Koç University Research Center for Translational Medicine (KUTTAM), funded by the Presidency of Turkey, Head of Strategy and Budget.

## Supplementary Data

### Supplementary Tables

**Table S1:**
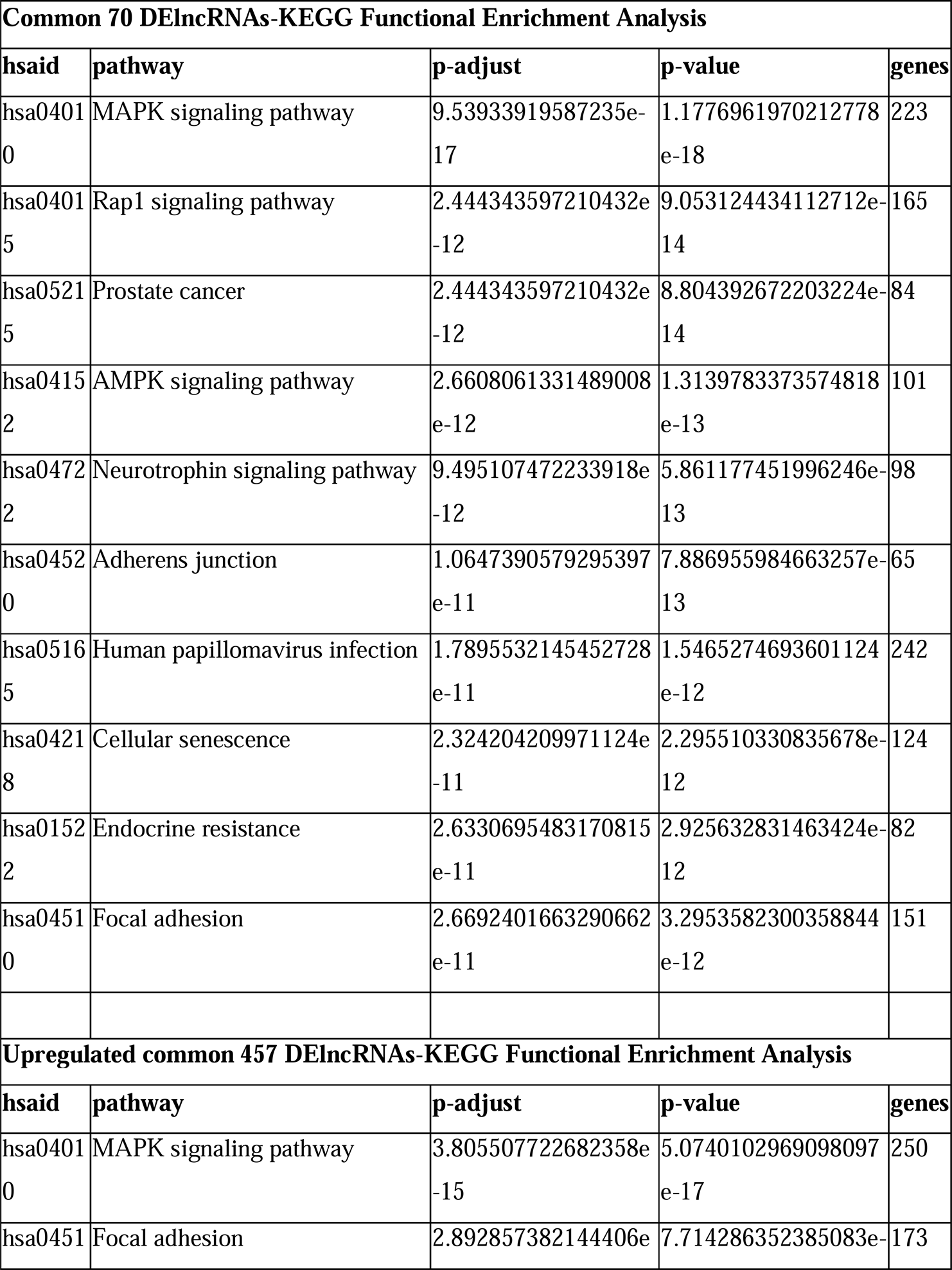
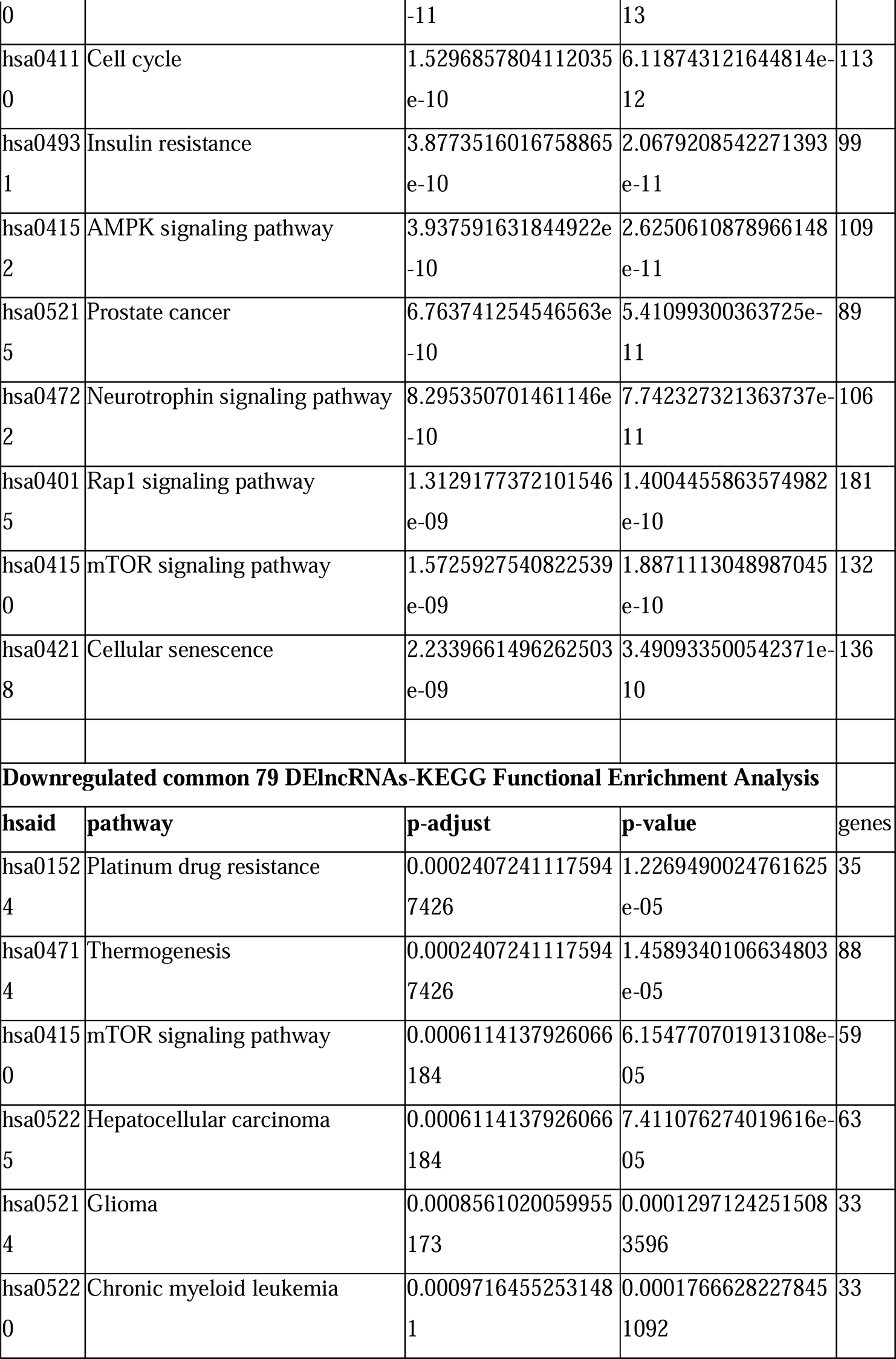
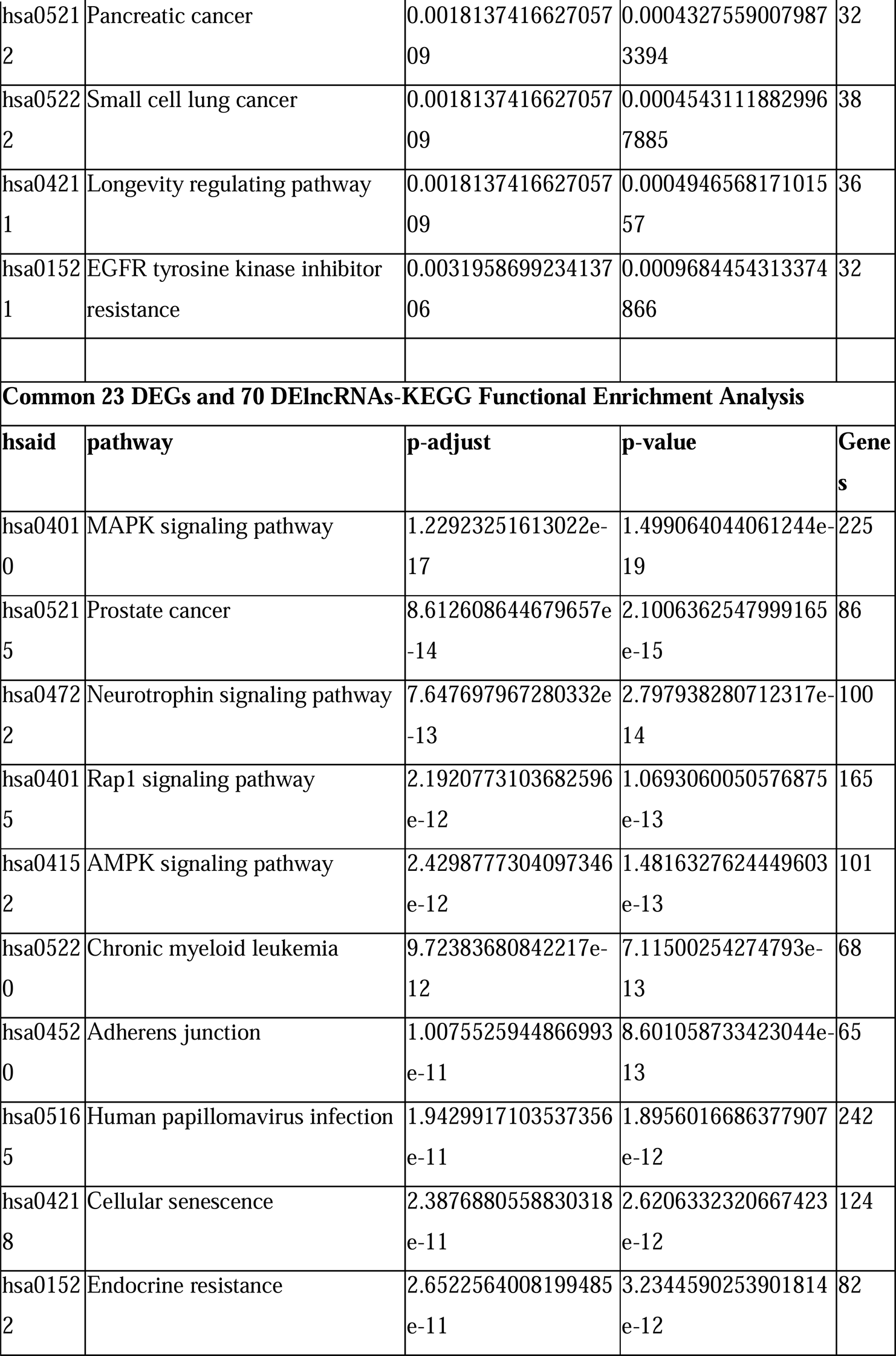
Top 10 functional enrichment (KEGG) analysis of upregulated, downregulated, common, and hub genes. Data were obtained from NCPATH database.

## Notes

### Competing Interest Statement

The authors have declared no competing interest.

### Author Declarations

Ethics committee of Cukurova University School of Medicine gave ethical approval for this work (Approval number: 356/22.05.2021)

