## Supplementary figures and images for "Comparative transcriptomic analyses of peripheral blood mononuclear cells of patients with non-pneumonia and severe pneumonia at 1 year-Long-COVID-19"

### Figure S1

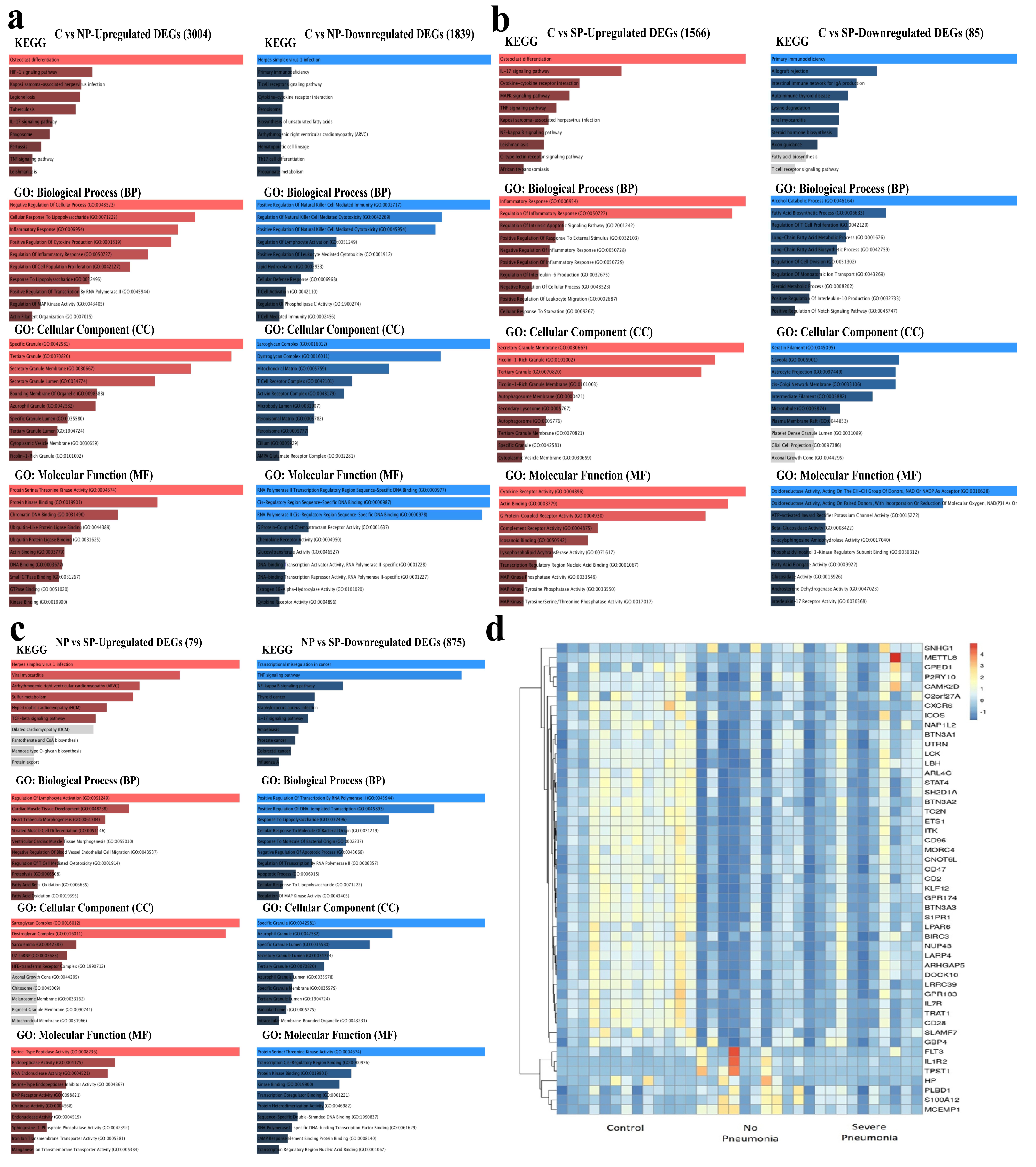

### Figure S2

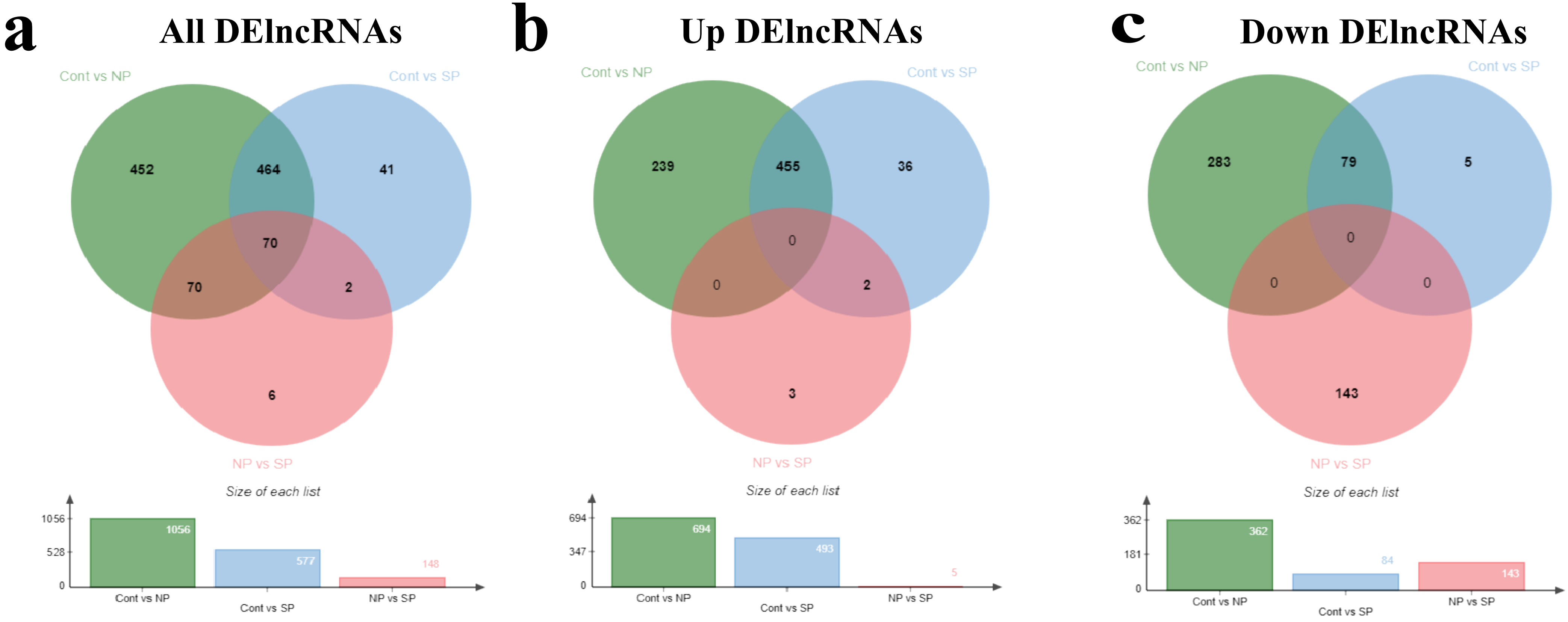
